# Co-Developing a Women-Centered HIV Prevention Intervention to Reduce Stigma, Increase HIV Self-Testing, and Improve Pre-Exposure Prophylaxis (PrEP) Uptake in Ghana (WISE WOMAN): A Study Protocol

**DOI:** 10.64898/2026.04.01.26349993

**Authors:** Gloria Aidoo-Frimpong, Yaa Adutwumwaa Obeng, Abass Tando Abubakar, Winfred Kofi Mensah, Daniel Selase Anyidoho

**Affiliations:** Department of Epidemiology and Environmental Health, School of Public Health and Professions, University at Buffalo, State University of New York, Buffalo, NY, USA; Center for Interdisciplinary Research on AIDS, Yale University, New Haven, CT, USA; School of Public Health, Ensign Global University, Akosombo, Ghana; Department of Health Information, St. Patrick Catholic Hospital, Offinso, Ghana; Department of Medical Statistics, Hawa Memorial Saviour Hospital, Ghana; Department of Laboratory Services, Hawa Memorial Saviour Hospital, Ghana; Transforming the HIV Response through Innovation and Equity (THRIVE) Lab at UB, Buffalo, New York, U.S.A; Department of Health Services, Brigham Young University, Idaho, USA

**Author notes:** **Funding:** UB’s Offices of International Education (OIE) and Vice President for Research and Economic Development (OVPRED) Seed Funding for External Grants in Global and International Research. **Competing Interest:** The authors have no competing interests. **Data availability:** No datasets were generated or analyzed during the current study, as this manuscript describes a study protocol. Data generated from the pilot implementation study will not be publicly available due to ethical and privacy considerations related to the sensitive nature of the data and the need to protect participant confidentiality. De-identified data may be made available upon reasonable request to the corresponding author, subject to institutional review board approval and applicable data sharing agreements.

**Keywords:** HIV prevention, adolescent girls and young women, HIV self-testing, pre-exposure prophylaxis, community-based participatory research, human-centered design, digital health, WhatsApp, Ghana, women-centered care

## Abstract

**Background:** Young women in Ghana (18–35 years) remain disproportionately affected by HIV due to intersecting structural and social challenges, including stigma, gendered power dynamics, and limited access to women-centered prevention services. Although HIV self-testing (HIVST) and pre-exposure prophylaxis (PrEP) are effective biomedical prevention strategies, uptake among young Ghanaian women remains low. Barriers include limited awareness, persistent stigma, and a lack of culturally relevant, youth-responsive prevention approaches. The WISE WOMAN study aims to address these gaps by developing and piloting a women-centered HIV prevention intervention co-created with young women in Ghana.

**Methods:** This protocol describes a pilot implementation study of a women-centered HIV prevention intervention that will be delivered via WhatsApp. The intervention is informed by community-based participatory research and human-centered design approaches to enhance cultural relevance and responsiveness to young women’s lived experiences. The study will enroll 50 young women aged 18–35 years who will participate in a four-week WhatsApp-based intervention designed to increase HIV prevention knowledge, reduce stigma, and support engagement with HIVST and PrEP. Implementation outcomes, including feasibility, acceptability, and appropriateness, will be assessed using mixed methods. Quantitative data will be collected through baseline and post-intervention surveys, including the PIERS-22 engagement scale, and will be analyzed using descriptive statistics and paired comparisons. Qualitative data from group interactions and post-intervention interviews will be analyzed using thematic analysis. The study has received ethical approval from the University at Buffalo Institutional Review Board (STUDY00009328) and the Ensign Global College Ethics Committee (IRB/EL/AF-02/2025) and is registered at ClinicalTrials.gov (NCT07003789).

**Discussion:** This protocol outlines the design and methods for a digitally delivered, women-centered HIV prevention intervention grounded in participatory approaches. The planned pilot study will generate critical implementation evidence on the feasibility, acceptability, and appropriateness of a WhatsApp-based, co-designed intervention, informing future adaptation, scale-up, and integration of culturally grounded HIV prevention strategies for young women in Ghana and similar settings.

## Introduction

Adolescent girls and young women (AGYW) in sub-Saharan Africa continue to be disproportionately affected by HIV. In 2022, women and girls accounted for 53% of all people living with HIV globally and 46% of new infections [1]. In Sub-Saharan Africa (SSA), this proportion rose to 63%, with over 3100 AGYW aged 15-24 acquiring HIV every week [1]. In Ghana, gender disparities remain pronounced. As of 2024, 334,721 people were living with HIV; 68.5% were women and girls aged 15 and above, who also accounted for 67.4% of new infections and 56.8% of AIDS-related deaths [2]. These figures underscore the urgent need for tailored HIV prevention interventions for this demographic. These disparities are largely driven by intersecting structural and social factors, including economic dependency, unequal power dynamics, entrenched gender norms, and stigma surrounding HIV [3], [4], [5], [6]. Such conditions limit access to and engagement with HIV prevention services, particularly among AGYW. Despite being identified as a priority population, young women in Ghana continue to face significant barriers to using existing biomedical tools such as HIV self-testing (HIVST) and pre-exposure prophylaxis (PrEP).

HIVST presents a unique opportunity for young women to overcome stigma, ensure privacy, and enhance autonomy in HIV prevention. Yet despite its promise, awareness and use of HIVST among young Ghanaian women remains low, only 18.2% of women aged 15-24 have heard of HIVST, and just 2.4% had ever self-tested [7], [8]. Moreover, evidence suggests that structural barriers, such as economic dependency, unequal power dynamics, and entrenched gender norms, serve as critical structural barriers to the uptake of HIVST among women [9], [10], [11].

Similarly, PrEP offers another highly effective HIV prevention option, allowing women to protect themselves independently of male partner cooperation. A Ghana-specific study indicated that women were more likely than men to take PrEP when available [12]. However, PrEP awareness and uptake remain limited among Ghanaian women [13], with low uptake also documented among female sex workers [14]. This implementation gap, between progressive national policies and real-world uptake, highlights a critical disconnect between policy and practice.

Importantly, these shortfalls reflect missed opportunities rather than a lack of interest or need. Ghanaian women have demonstrated interest in PrEP when it is available [12], but persistent stigma, misinformation, and the absence of women-centered communication platforms continue to impede uptake [9], [10], [11], [13]. Thus, there is an urgent need for interventions that not only disseminate information but also resonate with young women’s experiences, preferences, and communication norms.

To address these challenges, the World Health Organization (WHO) has endorsed both HIVST and PrEP as effective tools for HIV prevention, especially for populations with limited access to traditional testing and prevention services [15], [16], [17], [18]. Ghana has taken steps to align with these recommendations by launching a national HIVST pilot (2021–2023) that distributed over 200,000 kits and by releasing national PrEP guidelines (initially in 2020, updated in 2023) [18], [19], [20]. However, these policy advances have not yet translated into widespread awareness or consistent uptake among young Ghanaian women [7], highlighting a persistent implementation gap. This disconnect between national guidelines and day-to-day use underscores the urgent need for women-centered approaches that can effectively operationalize policy on HIVST and PrEP into meaningful action.

Participatory methodologies such as community-based participatory research (CBPR) [21] and human-centered design (HCD) [22] have been effective in developing culturally grounded and acceptable health interventions. These approaches engage end-users throughout the design process, increasing ownership, cultural alignment, and sustainability [21], [22], [23], [24]. In Ghana, participatory approaches have enhanced the relevance of mental health and HIV interventions alike [25], [26], with evidence showing that co-creation strengthens engagement and feasibility The WISE Woman Study builds on this evidence by integrating CBPR and HCD to co-create a women-centered HIV prevention intervention with young women in Ghana. Prior to the pilot intervention, a formative co-creation workshop engaged young women and community stakeholders to identify barriers to HIVST and PrEP uptake and to inform the development of the intervention. Key insights from this formative stage highlighted the central role of relationship power dynamics, pervasive stigma surrounding HIV and sexuality, limited access to trusted and youth-friendly sexual health information, and a strong preference for private, peer-supported platforms for discussing HIV prevention. In response, participants co-designed a WhatsApp-delivered intervention that emphasizes privacy, relatability, peer engagement, and actionable guidance on HIVST and PrEP. By embedding participatory design within a digitally delivered intervention model, the WISE Woman Study advances a novel approach to HIV prevention that integrates user-centered design, peer support, and scalable mobile health delivery. Accordingly, this protocol outlines the rationale, intervention architecture, study design, analytic strategy, and ethical framework for evaluating the feasibility, acceptability, and preliminary impact of this approach. Findings from this study will generate critical implementation evidence to inform the design, adaptation, and scale-up of women-centered HIV prevention strategies in Ghana and similar low-resource settings.

## Method

### Study Design

This study will be a pilot implementation study evaluating a women-centered HIV prevention intervention delivered via WhatsApp to young women in Ghana. The primary objective of the pilot will be to assess key implementation outcomes, including feasibility, acceptability, and appropriateness of the intervention in a real-world, digitally mediated setting. Secondary objectives will include examining preliminary changes in HIV prevention knowledge, perceived stigma, and willingness to use HIVST and PrEP. A mixed-method pre-post design will be employed. Quantitative data will be collected through baseline and post-intervention surveys, while qualitative data will be obtained through post-intervention interviews to capture participant experiences, perceived value, and recommendations for improvement.

### Ethical Considerations

This study has been reviewed and approved by two ethics committees. Ethical approval in the United States was obtained from the University at Buffalo Institutional Review Board (STUDY00009328; approved July 2025), which determined that the study involves no greater than minimal risk and is subject to continuing review, with required reporting within 30 days of study closure. Ethical approval in Ghana was obtained from the Ensign Global College Ethics Review Committee (IRB/EL/AF-02/2025; approved May 2025). All study procedures will be conducted in accordance with applicable ethical guidelines and regulations in both countries. The full IRB-approved study protocol is provided as Supporting Information (see S1 File). This study protocol is registered at ClinicalTrials.gov (NCT07003789; https://clinicaltrials.gov/study/NCT07003789). All participants will provide verbal informed consent prior to enrollment using IRB-approved digital procedures. Following eligibility screening, participants will receive the consent form individually via WhatsApp to allow for private review. Participants will be encouraged to take sufficient time to review the document, ask questions via private messaging or phone calls, and proceed only if they feel comfortable. Verbal consent will be documented through verbal confirmation via secure messaging, including voice note confirmation, consistent with IRB-approved protocols, at least two weeks prior to participation in study activities. A single reminder will be sent if no response is received, with no further follow-up to avoid undue pressure. Participants will be informed of their right to withdraw at any time without penalty.

The SPIRIT recommended schedule of enrollment, intervention, and assessment for participants is presented in Fig 1 [28]. This includes enrollment procedures (such as screening and consent), intervention activities, implementation measures, and quantitative and qualitative assessments across study time points.

**Fig 1.**
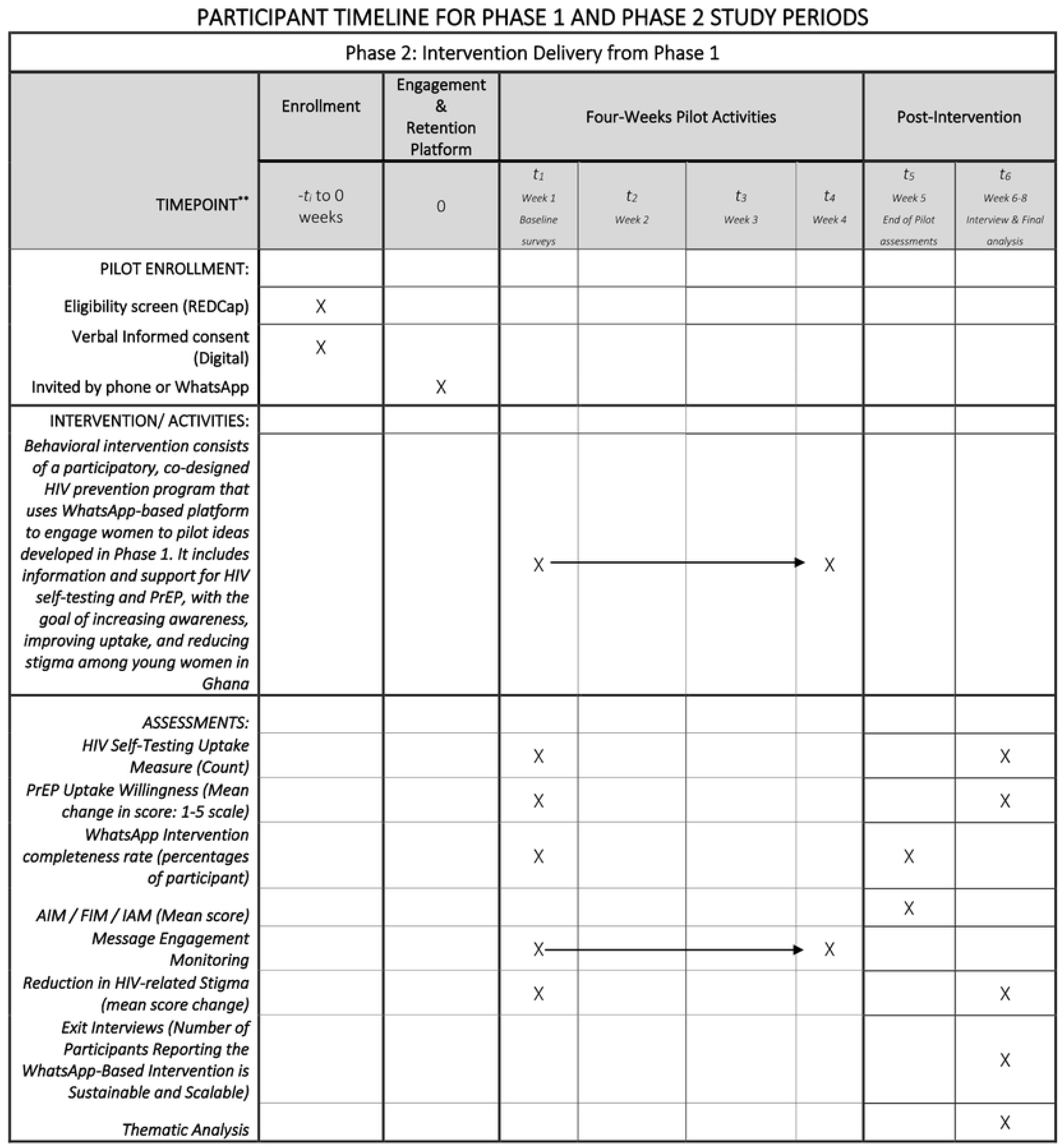
SPIRIT schedule of enrolment.

The total target sample size pilot study is 50 participants. An estimated 100 individuals will be screened to enroll the final sample, corresponding to an anticipated screening-to-enrollment ratio of approximately 2:1. This approach ensures adequate recruitment while minimizing participant burden during early-stage implementation research. As a pilot implementation study, the trial is not powered to detect effectiveness outcomes; however, findings from this study will inform the design and sample size calculations for a future fully powered effectiveness trial.

### Study Status and Timeline

At the time of manuscript submission, the study has not yet commenced participant recruitment or data collection. The study is currently in the preparatory phase, including finalization of intervention materials and study procedures. Participant recruitment is expected to begin in May 2026, with intervention implementation planned for August 2026. Post-intervention surveys and qualitative exit interviews are expected to be completed by October 2026. Data analysis will follow completion of data collection and is expected to conclude by December 2026. Final study results are anticipated by March 2027.

### Participants and Recruitment

#### Participants

We will enroll 50 young women (ages 18–35) who must reside in Greater Accra or Eastern Region, own a smartphone with WhatsApp capabilities, and feel comfortable discussing sexual/reproductive health. All participants must be able to provide verbal informed consent. Exclusion criteria include any cognitive impairment that would prevent participation. Community stakeholders will not participate in the pilot phase. The target sample of 50 participants is appropriate for a pilot implementation study focused on assessing acceptability, feasibility, appropriateness, engagement, and preliminary behavioral signals rather than effectiveness outcomes, consistent with guidance that pilot studies are not intended to be powered for hypothesis testing [40], [41], [42]. Pilot studies of digital and mobile health interventions commonly enroll 40–60 participants to provide stable estimates of implementation outcomes and to inform refinement prior to larger-scale trials [40], [41], [43], [44]. To reach the enrollment target, approximately 100 individuals will be screened, accounting for eligibility screening, consent refusal, and anticipated attrition.

#### Recruitment

Recruitment announcements (via WhatsApp, clinics, women’s centers) will invite young women to a screening survey. Enrollment will be on a first-come, first-eligible basis, with an emphasis on diversity in age and background. All participants must consent (via an electronic consent) before beginning the intervention (e.g., sending a verbal consent and receiving a voice/text consent confirmation). We will collect baseline demographic data to describe the sample. Efforts will ensure women from both regions are represented.

#### Compensation

Participants enrolled in the pilot intervention will be eligible to receive GH₵300 (approximately $30) upon completion of all required study activities, including participation in the four-week intervention and completion of both baseline and post-intervention surveys. Participants who additionally take part in qualitative exit interviews will receive an extra GH₵100 (approximately $10). Participants may withdraw from the study at any time without penalty

#### Confidentiality and Data Protection

Personal information about potential and enrolled participants will be collected, managed, and protected in accordance with ethical guidelines and IRB requirements to ensure confidentiality throughout all stages of the study.

**Before enrollment**, limited personal information (e.g., phone number and eligibility responses) will be collected through a secure, password-protected REDCap screening survey. Access to screening data will be restricted to authorized study personnel only. Individuals who do not meet eligibility criteria will not be retained in study records beyond what is necessary for screening documentation.

**During the study**, enrolled participants will be assigned unique study identification numbers, and all survey and qualitative data will be de-identified prior to analysis. Identifiable information (e.g., phone numbers used for WhatsApp communication) will be stored separately from study data in encrypted, password-protected files. The WhatsApp group will be configured to protect participant privacy, including the use of study-specific guidelines (e.g., use of nicknames, prohibition of message forwarding or screenshots, and encouragement of private communication with facilitators for sensitive topics). Participants will be informed of potential privacy risks associated with group-based digital platforms and will have the option to engage at their comfort level.

**Data sharing among the research team** will be limited to de-identified datasets. Only authorized team members will have access to identifiable information for purposes of study coordination and participant communication.

**After study completion**, all identifiable data will be securely stored on encrypted, password-protected institutional servers for a period consistent with IRB requirements. Any data used for dissemination will be fully de-identified and reported in aggregate form to prevent identification of individual participants. No personally identifiable information will be shared in publications or presentations.

These procedures are designed to minimize risks related to confidentiality and ensure that participant privacy is protected before, during, and after participation in the study.

#### Intervention Delivery

The pilot intervention will be delivered over four weeks through a structured, group-based WhatsApp platform. WhatsApp is widely used among young women in Ghana and supports multimedia communication, making it an appropriate and scalable platform for delivering HIV prevention interventions. Participants will be enrolled in a closed WhatsApp group moderated by trained facilitators. The group will serve as the primary platform for delivering interventions, engaging participants, and fostering peer interaction. Participants will receive standardized content according to a predefined weekly schedule, and they will also have the option to communicate privately with facilitators about sensitive questions or concerns. The intervention will be guided by a co-designed framework developed with young women, which emphasizes the importance of privacy, relatability, peer engagement, and practical, action-oriented learning. The intervention will include three integrated components:

#### 1. Structured Content Delivery

Participants will receive three multimedia content modules per week (e.g., short videos, audio messages, and visual materials). Content will address HIV prevention topics, including HIVST, PrEP, stigma, relationship dynamics, and access to services (Table 1).

**Table 1:** Weekly Content Calendar for the 4-Week WhatsApp Pilot Intervention.

| Week | Weekly Goal | Day | Content Type | Description |
| --- | --- | --- | --- | --- |
| **Week 1:**<br>**Foundations and Awareness** | Introduce HIV, HIVST, and PrEP in relatable ways that reduce fear and build trust | Mon | Video 1 | Welcome message; group rules; baseline survey reminder |
|  |  | Wed | Video 2 + Prompt | “When you hear self-testing, what comes to mind?” |
|  |  | Fri | Video 3 + Poll | “Before today, had you heard of PrEP?” (Yes/No) |
|  |  | Weekend | Soft Engagement | Reflection: “What stood out to you this week?” |
| **Week 2: Stigma, Relationships, and Social Realities** | Surface lived barriers while validating women's experiences | Mon | Video 4 + Prompt | "What would make you feel safe when seeking care?" |
|  |  | Wed | Video 5 + Poll | "Can a partner's reaction influence testing or PrEP decisions?" (Yes/No) |
|  |  | Fri | Video 6 + Prompt | "What change in your community would help women most?" |
|  |  | Weekend | Engagement | Appreciation message; anonymous voice note option |
| **Week 3: Practical Skills and Decision-Making** | Build confidence in HIVST use and PrEP access | Mon | Video 7 + Poll | "Do you feel more confident using a test after watching this?" |
|  |  | Wed | Video 8 + Prompt | "What would make the process easier for you?" |
|  |  | Fri | Video 9 + Q&A | PrEP visit expectations; open Q&A ("No question is too small") |
|  |  | Weekend | Support Message | Optional self-testing support and guidance |
| **Week 4: Strength, Autonomy, and Future Planning** | Support self-efficacy and future HIV prevention behaviors | Mon | Video 10 + Prompt | “What strengths help you care for your health?” |
|  |  | Wed | Video 11 + Poll | “Would you test again in the next 6 months?” |
|  |  | Fri | Video 12 + Reflection | “What changed for you over these 4 weeks?” |
|  |  | Weekend | Closing | Post-survey reminder and thank-you message |

#### 2. Interactive Engagement

Participants will engage in guided discussions, polls, and reflection prompts designed to reinforce learning and encourage peer-to-peer interaction (Table 1). Facilitators will actively moderate discussions, respond to questions, and correct misinformation.

#### 3. Activity-Based Behavioral Tasks

Participants will be invited to complete structured weekly activities designed to translate knowledge into real-world action. Activities will focus on building awareness, communication skills, health system navigation, and future prevention planning (Table 2). Participation in activities will be encouraged but voluntary.

**Table 2.**
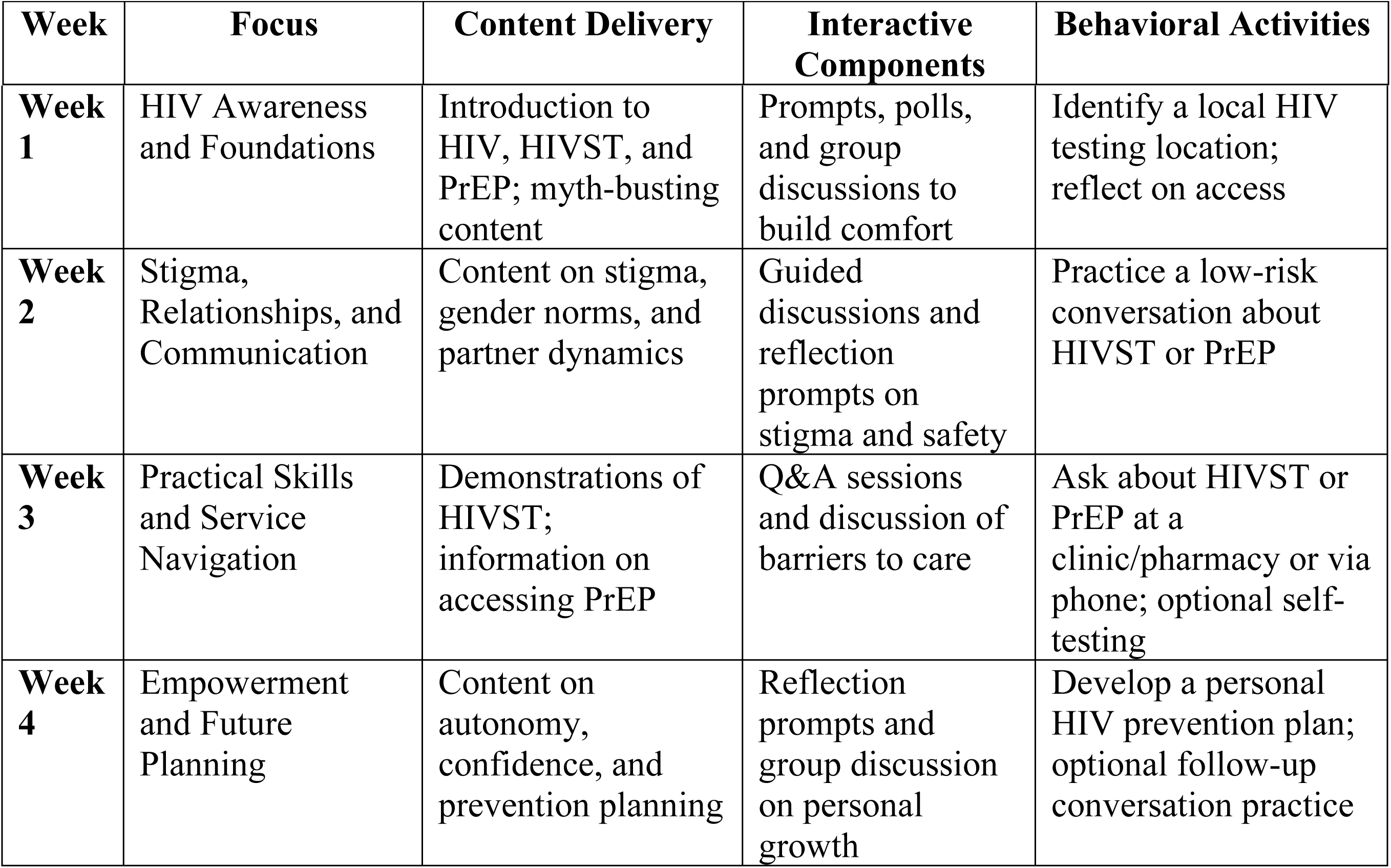
Weekly Intervention Structure and Content.

| Week | Focus | Content Delivery | Interactive Components | Behavioral Activities |
| --- | --- | --- | --- | --- |
| **Week 1** | HIV Awareness and Foundations | Introduction to HIV, HIVST, and PrEP; myth-busting content | Prompts, polls, and group discussions to build comfort | Identify a local HIV testing location; reflect on access |
| **Week 2** | Stigma, Relationships, and Communication | Content on stigma, gender norms, and partner dynamics | Guided discussions and reflection prompts on stigma and safety | Practice a low-risk conversation about HIVST or PrEP |
| **Week 3** | Practical Skills and Service Navigation | Demonstrations of HIVST; information on accessing PrEP | Q&A sessions and discussion of barriers to care | Ask about HIVST or PrEP at a clinic/pharmacy or via phone; optional self-testing |
| **Week 4** | Empowerment and Future Planning | Content on autonomy, confidence, and prevention planning | Reflection prompts and group discussion on personal growth | Develop a personal HIV prevention plan; optional follow-up conversation practice |

## Data Collection and Analysis

### Quantitative Data

Research Assistants will administer online surveys (via REDCap) at baseline (Week 0) and post-intervention (Week 5). These will assess:

- *HIV Prevention Knowledge and Attitudes:* Questions on HIVST and PrEP awareness and stigma.
- *Acceptability, Feasibility, Appropriateness:* Using the validated 4-item Acceptability of Intervention Measure (AIM), Feasibility of Intervention Measure (FIM), and Intervention Appropriateness Measure (IAM) [35], [36], [47]. Each item is rated 1 (completely disagree) to 5 (completely agree); higher scale scores indicate greater acceptability, feasibility, or appropriateness [35], [36], [47]. We will calculate mean scores (±SD) for each measure.
- *HIVST Uptake:* Self-reported number of HIV self-tests completed during the study (participants can send a WhatsApp photo of the test as confirmation).
- *PrEP Willingness:* Using a 5-point Likert item (e.g. “I am likely to take PrEP in the next 3 months”), we will report mean change in intention.
- *Intervention Completion:* Percent of participants who complete all required activities; reading messages, participating in discussions, and both surveys.

### Qualitative Data

Semi-structured exit interviews will be conducted by the Research Assistants via WhatsApp voice call or in-person (depending on preference) with a purposive subsample of approximately 15–20 participants after the pilot (Week 6-8). Interviews will explore participants’ experiences (e.g. what they found useful, suggestions, perceived barriers/facilitators to using WhatsApp for HIV information). Interviews will be audio-recorded and transcribed.

### Analysis

Quantitative outcomes will be summarized descriptively. Mean scores on the AIM, FIM, and IAM scales will indicate overall acceptability, feasibility, and appropriateness [35], [36], [47]. we expect higher scores to reflect positive reception [36] Qualitative interview data will be analyzed thematically (Braun & Clarke) [39]. We will code transcripts inductively, identify themes related to user experience, perceived relevance, and sustainability of the WhatsApp intervention. Notably, we will map themes onto Proctor’s implementation outcomes (acceptability, feasibility, appropriateness [36] to enrich quantitative findings with participant context. For example, if AIM scores are high, interviews may reveal which aspects (e.g. message tone, peer support) drove acceptability. Across both phases, findings will be triangulated: Phase 1 outputs directly shape Phase 2 content, and Phase 2 evaluation will inform the viability of the co-designed intervention. This iterative process aligns with implementation research best practices, ensuring the final program is grounded in user input and evidence [27], [36]. R will be employed as the statistical software.

### Dissemination

All data will be de-identified and stored securely on password-protected servers accessible only to the research team. Audio recordings, transcripts, and WhatsApp interactions will be anonymized before analysis. Participants may use pseudonyms during WhatsApp group activities, and privacy guidelines will be communicated clearly throughout the study. Upon completion of the study, study findings will be disseminated through peer-reviewed journal articles, conference presentations, policy briefs, community feedback sessions, and reports shared with local stakeholders and participating organizations. Results will also be communicated to participants through WhatsApp and community outreach channels to support broader awareness and engagement in HIV prevention. Due to the sensitive nature of the data, individual participant data (IPD) will remain confidential and will not be shared publicly to protect confidentiality.

### Expected Outcomes

The pilot study will assess key implementation outcomes, including feasibility, acceptability, and appropriateness of the intervention, as well as preliminary behavioral and psychosocial indicators relevant to HIV prevention. Specifically, the study will examine participant engagement, perceived acceptability and feasibility of WhatsApp-based delivery, and changes in HIV prevention knowledge, stigma, and willingness to use HIV self-testing and PrEP. Qualitative data will explore participants’ experiences, the perceived relevance of the intervention content, and recommendations for improving delivery and scalability. Findings from this pilot will inform intervention refinement and the design of a future fully powered effectiveness trial. Expected outcomes of the pilot study are summarized in Table 3.

**Table 3.** Expected Outcomes of Phase 2 Pilot Intervention.

| **Outcome Category** | **Expected Outcome** |
| --- | --- |
| **Implementation Outcomes** | • High scores on Acceptability (AIM), Feasibility (FIM), and Appropriateness (IAM). (Scales. AIM/FIM/IAM scores $\geq 4.0$ on average (out of 5) may be considered evidence of high acceptability/feasibility)<br>• $\geq 80\%$ intervention completion rate across 4 weeks.<br>• Strong message engagement (e.g., active participation in chat prompts, audio feedback, etc.). |
| **HIV Prevention Outcomes** | • Improved HIV prevention knowledge (pre/post comparison).<br>• Reduced HIV-related stigma (internalized and anticipated).<br>• Uptake of HIVST during the pilot (verified via photo or report).<br>• Increased willingness to use PrEP. |
| **Qualitative Outcomes** | • Thematic insights into participant experience with intervention delivery.<br>• Feedback on intervention sustainability, relevance, and scalability.<br>• Qualitative data aligned with Proctor’s framework to complement implementation metrics. |

### Provisions to Monitor the Data to Ensure the Safety of Subjects

The research team will implement a structured plan to periodically assess the collected data to ensure participant safety and monitor potential harm and benefits. This includes:

The study team will conduct biweekly reviews of participant survey responses and interview transcripts to identify emerging patterns related to distress, stigma, or other concerns. Direct check-ins will be scheduled every two weeks with participants through WhatsApp or in-person meetings to assess their well-being and address any concerns. If risks such as heightened distress, safety concerns, or increased stigma are identified, appropriate mitigation strategies will be implemented, including referrals to support services and modifications to study materials.

## Discussion

The WISE Woman Study protocol will address persistent disparities in HIV prevention among AGYW in Ghana by integrating a participatory, women-centered design approach with digital delivery. This is particularly relevant in sub-Saharan Africa, where AGYW continue to bear a disproportionate burden of the HIV epidemic. Structural barriers: stigma, unequal gender dynamics, and limited access to women-friendly services, hindering prevention strategies such as HIVST and PrEP [3], [7]. Notably, previous studies in Ghana have documented critically low levels of HIVST awareness and uptake [7], [8]. At the same time, stigmatizing attitudes toward people living with HIV remain high among young women in Ghana, and stigma has been shown to significantly reduce testing uptake [3], [48]. These gaps underscore an implementation lag between policy and practice, particularly for AGYW.

This protocol directly addresses this implementation gap by piloting a women-centered HIV prevention intervention delivered through WhatsApp, a widely used and accessible digital platform that has demonstrated potential for supporting privacy, acceptability, and engagement in digital health interventions across Africa[46], [49]. The intervention was informed by formative work conducted with young women and community stakeholders, which helped identify key contextual barriers to HIV prevention and informed the design, tone, and delivery format of the digital intervention. By incorporating CBPR and HCD, the study is expected to yield an intervention that reflects women’s priorities, lived experiences, and communication preferences. Subsequently, these approaches have been associated with improved contextual relevance and increased the likelihood of adoption and sustainability in prior participatory design studies in Ghana and Kenya [21], [22], [25], [34].

A key strength of this protocol is its methodological rigor and real-world applicability. The pilot implementation study will evaluate core implementation outcomes, including high acceptability, feasibility, and appropriateness, as measured using validated implementation science instruments (AIM, FIM, IAM) [35], [36]. These measures are widely used in early-stage intervention research and are recommended for assessing implementation success prior to scale-up [35], [36]. Furthermore, high intervention completion and message engagement rates during the four-week pilot would further support the feasibility of WhatsApp-based delivery, consistent with evidence demonstrating the effectiveness of mobile messaging platforms for delivering HIV and sexual health interventions to AGYW in low-resource settings [46], [51].

In addition to implementation outcomes, the study anticipates short-term behavioral and psychosocial outcomes, including improved HIV prevention knowledge, reduced internalized and anticipated stigma, uptake of HIVST during the pilot period, and increased willingness to consider PrEP use. These proximal outcomes are theoretically linked to future prevention behaviors and are commonly used indicators in pilot HIV prevention studies [19], [20], [23]. Prior digital and peer-supported interventions in sub-Saharan Africa have demonstrated that women-centered, confidential, and culturally tailored messaging can improve HIV-related knowledge, reduce stigma, and increase testing behaviors among young women [23], [46], [48].

Despite these strengths, several implementation challenges may arise. First, digital access and connectivity may be uneven across participants. To mitigate this, the study will provide mobile data support and peer facilitator reminders, as well as asynchronous participation options. Second, stigma may persist even in digital spaces, potentially limiting engagement. This will be addressed through co-created messaging and trained peer educators to normalize discussions and maintain a supportive tone. A third concern is intervention fatigue or disengagement across the four-week pilot. To address this, content will be designed to be brief, engaging, and interactive, including audio and visual formats, consistent with evidence on digital health retention strategies among women [23]. This WISE Woman study protocol is expected to generate a scalable model for women-centered model for HIV prevention that integrates co-creation, digital delivery, and implementation science frameworks. By generating implementation-relevant data from a Ghanaian context, where women-focused HIVST and PrEP interventions remain limited, this protocol responds directly to calls for evidence that bridges the policy-to-practice gap in HIV prevention for AGYW in low-and middle-income countries (LMICs) [13], [14].

## Author Contributions

**Conceptualization, Methodology, Supervision:** Gloria Aidoo-Frimpong

**Writing – original draft:** Gloria Aidoo-Frimpong, Yaa Adutwumwaa Obeng, Abass Tando Abubakar, Winfred Kofi Mensah, Daniel Selase Anyidoho

Writing – review & editing: Gloria Aidoo-Frimpong, Yaa Adutwumwaa Obeng, Abass Tando Abubakar, Winfred Kofi Mensah, Daniel Selase Anyidoho

## Supporting Information

S1 Checklist. SPIRIT Checklist

S1 File. Detailed protocol submitted to ethics committee

S2 File. Verbal Consent Pilot

## Data Availability

No datasets were generated or analyzed during the current study, as this manuscript describes a study protocol. Data generated from the pilot implementation study will not be publicly available due to ethical and privacy considerations related to the sensitive nature of the data and the need to protect participant confidentiality. De-identified data may be made available upon reasonable request to the corresponding author, subject to institutional review board approval and applicable data sharing agreements.

